# Risk Stratification of Patients With Moderate Aortic Stenosis Using Aortic Valve Calcium

**DOI:** 10.1101/2025.08.01.25332711

**Authors:** HangYu Watson, Syed Ahmad, Kunal N. Patel, Philippe Pibarot, Jonathon Leipsic, Maan Awad, Sudarshan Balla, Irfan Zeb

## Abstract

**Objectives:** To evaluate whether aortic valve calcium (AVC) scoring by cardiac CT can enhance risk stratification and identify high-risk patients among those with moderate aortic stenosis (AS).

**Background:** Moderate AS is traditionally considered benign, yet emerging evidence suggests a subset may experience adverse outcomes similar to those with severe AS. AVC scoring has been validated for diagnosing severe AS but its prognostic value in moderate AS remains unclear.

**Methods:** A retrospective cohort study of 606 patients with moderate aortic stenosis (MAS) for whom aortic valve calcium (AVC) assessment was recommended on echocardiogram reports. Patients were categorized into two groups: those who underwent AVC (n=159) and those who did not (n=447). Primary outcomes included aortic valve replacement (AVR) and all-cause mortality. Secondary outcomes included time to severe AS diagnosis and time to AVR.

**Results:** A significantly higher proportion of patients in the CT cohort reached a diagnosis of severe AS (28.8% vs. 19.9%, p=0.037) and underwent valve intervention (32.6% vs. 14.1%, p<0.001) compared to the no-CT cohort. Patients underwent valve intervention mostly at the severe AS stage. All-cause mortality was markedly lower in the CT cohort (7.6% vs. 24.9%, p<0.001); difference being driven by excess non-cardiovascular mortality in the no-CT cohort (61.0% vs. 7.9%, p<0.001). Patients with severe AVC (males AVC >2000 AU and females >1200 AU) were more likely to undergo valve intervention compared with non-severe AVC (HR 2.60, 95% CI 1.42-4.74, p=0.002). Patients in the CT cohort were more likely to undergo valve intervention (HR 3.17, CI 2.04-4.91, p<0.001), and had a significantly lower all-cause mortality (HR 0.32, CI 0.16-0.61, p=0.001) compared to the no-CT cohort.

**Conclusion:** A practice algorithm incorporating AVC for risk stratification of MAS patients can lead to better outcomes. Role of AVC based decision-making in patients with MAS needs further study in a randomized trial.

## 1. Introduction

There has been an increasing focus on moderate aortic stenosis (MAS) due to its association with adverse cardiovascular events, including death. ^[1]^ The diagnosis is straightforward if the aortic valve area (AVA) and hemodynamic data findings are concordant (AVA 1.0-1.5 cm^2^, mean gradient 20-40 mmHg and aortic valve peak velocity [AV Vmax] 3-4 m/sec)].^[2]^ Diagnostic difficulties can arise when there is mismatch between AVA and pressure gradient, with hemodynamics falling in the mild severity (mean gradient <20 mmHg and Vmax <3 m/sec) and AVA in the moderate range (AVA 1.0-1.5 cm^2^) or hemodynamics in the moderate range (mean gradient 20-40 mmHg and Vmax 3-4 m/sec) and AVA in the severe range (<1.0 cm^2^) under normal flow conditions. Additional layers of complexity in the diagnosis of MAS can arise from measurement errors. These errors can arise from overestimation of left ventricular outflow tract (LVOT) diameter, improper alignment of the Doppler signal, overestimation of LVOT Doppler signals, lack of use of additional imaging windows, and/or inaccurate interpretation of complex hemodynamic data. Various measures are being utilized to risk stratify MAS patients such as myocardial fibrosis ^[3]^, abnormal global longitudinal strain ^[4]^ and myocardial systolic dysfunction. ^[5]^. A distinct subset of low-gradient AS patients have been identified that are characterized by a large LVOT with elevated AVC identified a severe AS phenotype despite an AVA_CT-LVOT_ >1.2 cm^2^, best characterized by indexed AVA and AVC. ^[6]^ Such patients with severe AVC had worse composite outcomes of all-cause mortality or valve intervention. ^[6]^ AVC can also be used to differentiate moderate from mild AS. ^[7]^ However, currently AVC assessment for risk stratification of MAS is not a routine clinical practice. To improve the diagnostic risk stratification and possible early referral for valve intervention for MAS patients, it was decided in 2021 to include a statement to consider AVC assessment if a diagnosis of MAS was reported on echocardiograms in our hospital system. The current study evaluates how this integration of AVC assessment for MAS evaluation changed the detection of severe AS and changed outcomes.

## 2. Method

### 2.1. Study population

The current study is a retrospective analysis of patients who underwent transthoracic echocardiography (TTE) at West Virginia University Hospital, displaying evidence of MAS between October 2021 and July 2024. The echocardiograms were interpreted by the local reading physicians. Once a diagnosis of MAS was made by the reading physician based on review of imaging and hemodynamic data, a reporting software pop up was included with the following statement: “consider ordering a CT Heart Calcium, to determine aortic valve calcium score for further risk stratification”. The reporting physician would include this statement in the final report if the diagnosis of MAS was made. The decision to order the AVC testing was left at the discretion of the treating physician who ordered the echocardiogram. The decision to perform AVC assessment was made after treating physician and patient discussion. For the current study, we selected patients that had a diagnosis of MAS on the reading physician reports and the statement for AVC testing was included in the final report. Additionally, to remove any discordant MAS categories, we included patients with aortic valve area (AVA) of 1.0-1.5 cm^2^, peak velocity of <4.0 m/sec and mean gradient of <40 mmHg. For the final cohorts, patients who had BAV between index TTE and the predefined outcomes or end of follow up period, and patients who had incomplete Doppler data for AS assessment on index TTE were excluded. These patients were followed over time via chart review until the predetermined endpoints were reached, which were valve intervention or all-cause mortality, at the end of the follow-up period.

Data collection encompassed downstream testing, including transthoracic and transesophageal echocardiography, low dose dobutamine stress echocardiography, cardiac catheterization, or heart team discussions. Patients suspected of moderate to severe AS (based on symptoms, hemodynamic data and AVC) were referred to the structural heart clinic for further evaluation and treatment decisions. Our structural heart team consists of an interventional cardiologist and a cardiac surgeon. Complex hemodynamic cases with unclear diagnosis were discussed in a heart team meeting involving multimodality imaging specialists, cardiac interventionalists, heart failure specialists, and cardiac surgeons. Patients underwent either surgical or percutaneous aortic valve intervention (SAVR or TAVR) once a diagnosis of severe AS was made by the heart team and in accordance with the published clinical guidelines. Patients were categorized into two groups: those who underwent AVC (CT cohort) and those who did not (no-CT cohort). The local Institutional Review Board (IRB) approved the research proposal, and the study complied with IRB regulations and HIPAA policies.

### 2.2. Transthoracic echocardiography

The acquisition of standard images followed the guidelines set forth by the American Society of Echocardiography (ASE). ^[8]^ The following parameters were collected from each index TTE report: left ventricular ejection fraction (LVEF), aortic valve (AV) peak velocity (Vmax), AV mean gradient, aortic valve area (AVA), stroke volume index (SVI) and AV dimensionless index (DI). Review of 2D images and hemodynamic data was done by the local reading cardiologist. For the current study analysis, a diagnosis of concordant moderate AS was made if the hemodynamics were consistent with AV mean gradient <40 mmHg, peak velocity <4 m/sec, and AVA 1.0-1.5 cm^2^.

### 2.3. Aortic valve calcium score

Patients underwent dedicated gated non-contrast CT scans on a 320-slice scanner (Aquilion Cannon Medical System^®^). The post-processed images were evaluated on a local workstation (Vitrea advanced visualization software, Vital imaging). AVC was measured on non-contrast CT scans using a previously published Agatston method. ^[9]^ Previously published AVC cutoffs were used for males and females to determine severe AS (males >2000 AU and females >1200 AU).^[8]^

### 2.4. Clinical outcomes

The outcome was defined as a composite of all-cause mortality and AV intervention (surgical or percutaneous valve replacement). The events were retrieved from electronic medical records. Follow up was calculated at death, valve intervention, or end of the study collection period. The first occurrence of any event (all-cause mortality or valve intervention) was considered as the outcome. The secondary outcome included median time to diagnosis of severe AS on follow-up imaging and median time to valve intervention. For the assessment of all-cause mortality, a meticulous review was conducted to ascertain whether mortality incidents were attributed to cardiac or non-cardiac causes by reviewing the death certification, hospice referral, or expiration summary.

### 2.5. Statistical analysis

Continuous data were presented as mean (± standard deviation) or median with interquartile range, and categorical data was presented as counts (%). We used non-parametric methods such as the Chi-squared test or Fisher’s exact test, where counts (%) were reported, and the Kruskal-Wallis test with Dunn-Bonferroni correction, where median [IQR] was reported. For mortality and aortic valve replacement outcomes, we constructed categorical models using the no-CT cohort as a reference and analyzed with Cox-proportional hazard regression to evaluate the impact of AVC scoring. Kaplan Meier curve analysis was performed for all-cause mortality and aortic valve replacement outcomes comparing CT and no-CT cohorts. All p-values were two-sided, with p<0.05 considered statistically significant. All statistical analyses were conducted with unweighted samples using R Studio (version 4.4.1).

## 3. Results

### 3.1. Baseline characteristics

The study included a total of 606 patients diagnosed with moderate aortic stenosis (AS) from index TTE reports that recommended consideration of AVC assessment. The final study population consisted of 409 patients that met the criteria of AVA 1.0-1.5 cm^2^, peak velocity <4 m/sec, and AV mean gradient <40 mmHg. Among these patients, 132 underwent AVC assessment (CT cohort), while 277 did not (no-CT cohort). There were no significant differences in baseline age or comorbidities between the CT– and no-CT cohorts, except for higher coronary artery disease (CAD) in the CT cohort (p=0.024) (Table 1). In the CT cohort, 41 patients were found to have severe AVC burden, representing 34% of the CT cohort. Overall, the median echocardiographic parameters were consistent with MAS in both cohorts (Table 1). AV peak velocity and mean gradient were higher in the CT-cohort compared to the no-CT cohort (p<0.001).

**Table 1.**
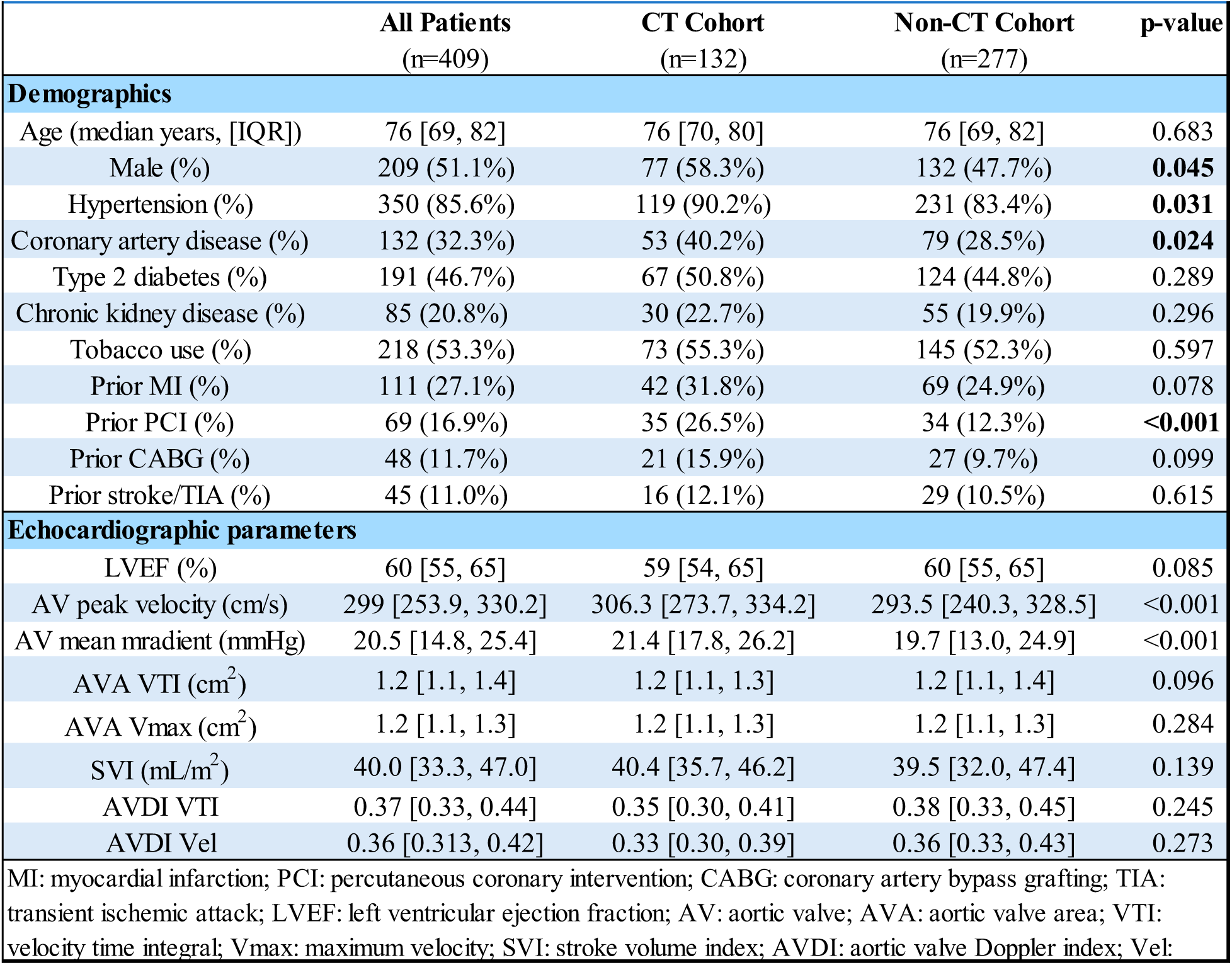
Baseline characteristics of moderate aortic stenosis patients who underwent cardiac CT for AVC assessment versus those who did not have cardiac CT.

### 3.2. Primary outcomes

Median follow up time for the full cohort was 564 days [IQR, 347-792], and was not significantly different between the CT cohort and no-CT cohort (p=0.104). Compared to the no-CT cohort, patients in the CT cohort were more likely to undergo AVR (HR 3.17, 95% CI 2.04-4.91, p<0.001; Figure 1). In the CT cohort, patients with severe AVC burden were more likely to undergo AVR compared with patients with non-severe AVC burden (HR 2.60, 95% CI 1.42-4.74, p=0.002; Figure 2). The AV hemodynamics, including AV peak velocity, AV mean gradient and AV DI, were worse among patients who underwent valve intervention compared to those who did not, in both CT– and no-CT cohorts. Median AVC was higher in the CT-cohort among patients who underwent AVR compared to those who did not receive valve intervention (1852 AU vs. 1051 AU, p<0.001). Similarly, the prevalence of severe AVC was also higher in patients who underwent AVR than those without valve intervention in the CT-cohort (48.8% vs. 24.1%, p=0.008).

**Figure 1.**
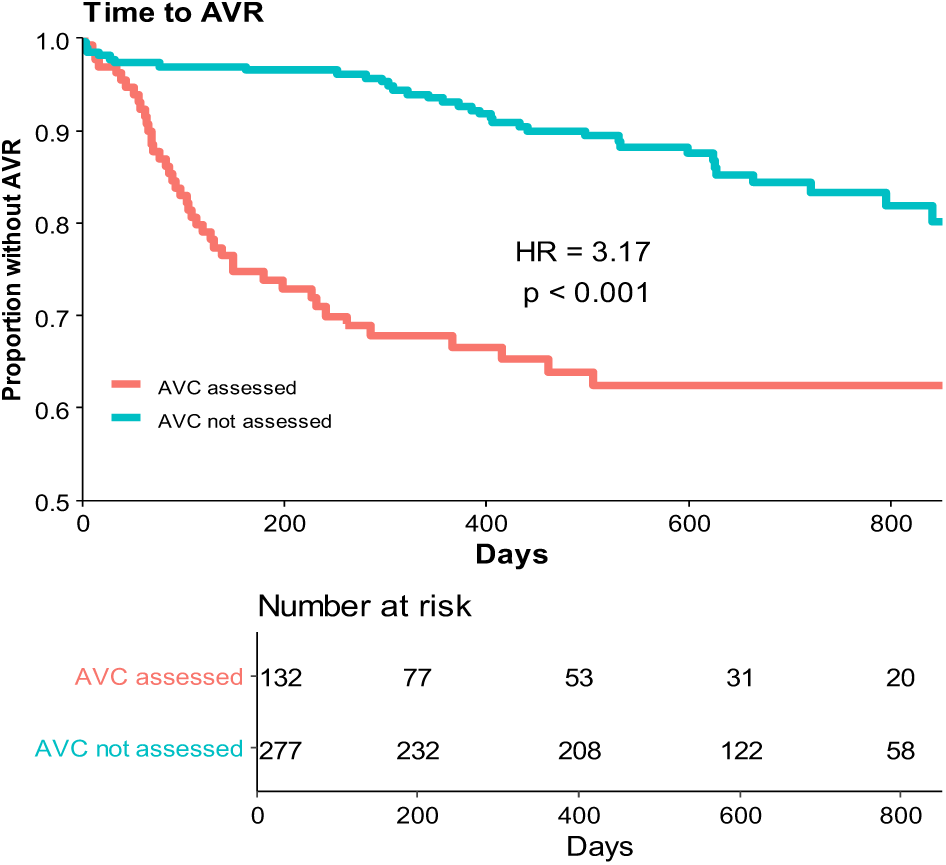
Kaplan-Meier Analysis of Time to Aortic Valve Replacement (AVR) by Aortic Valve Calcium (AVC) Assessment Status Kaplan-Meier curves comparing time to AVR in patients with moderate aortic stenosis who underwent computed tomography-derived AVC scoring (red line) versus those who did not (blue line). Patients in the AVC-assessed group had significantly earlier AVR compared to those not assessed (hazard ratio [HR] = 3.17; p < 0.001). The number at risk at each time point is shown below the plot.

**Figure 2.**
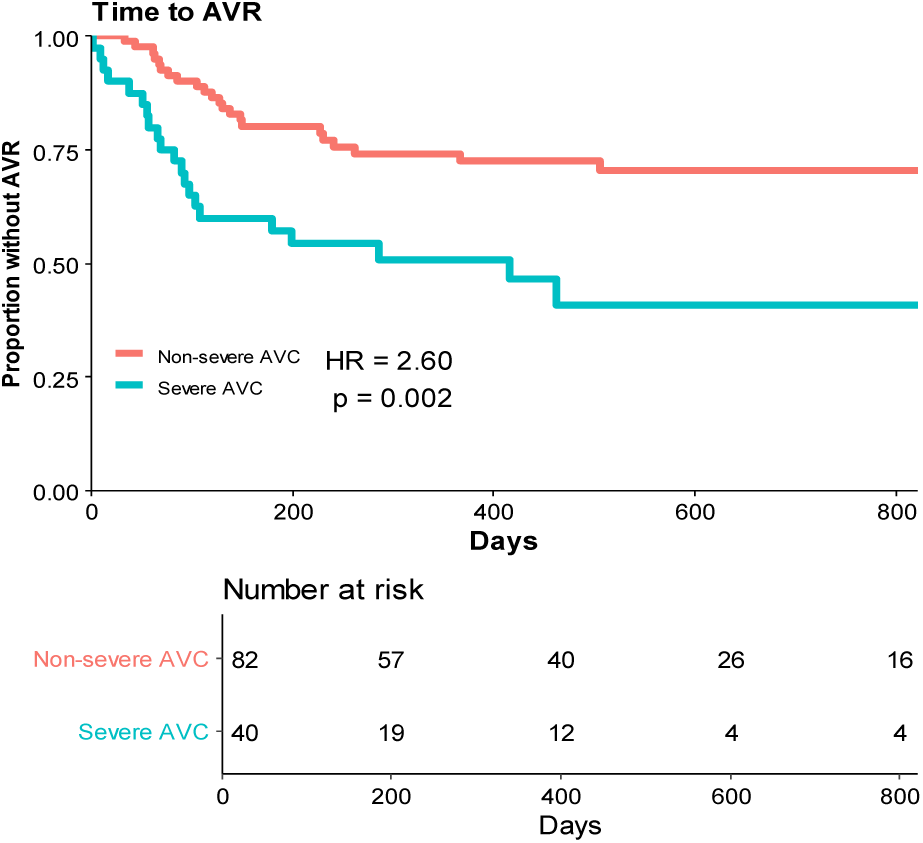
Kaplan-Meier Analysis of Time to Aortic Valve Replacement (AVR) by Severity of Aortic Valve Calcium (AVC) in the CT Cohort Kaplan-Meier curves showing time to AVR in patients with moderate aortic stenosis who underwent AVC scoring, stratified by severe versus non-severe AVC burden. Severe AVC was defined by established sex-specific thresholds (≥2000 AU for males and ≥1200 AU for females). Patients with severe AVC experienced significantly earlier AVR compared to those with non-severe AVC (HR = 2.60; p = 0.002). Number at risk for each group is displayed below the plot.

**Figure 3.**
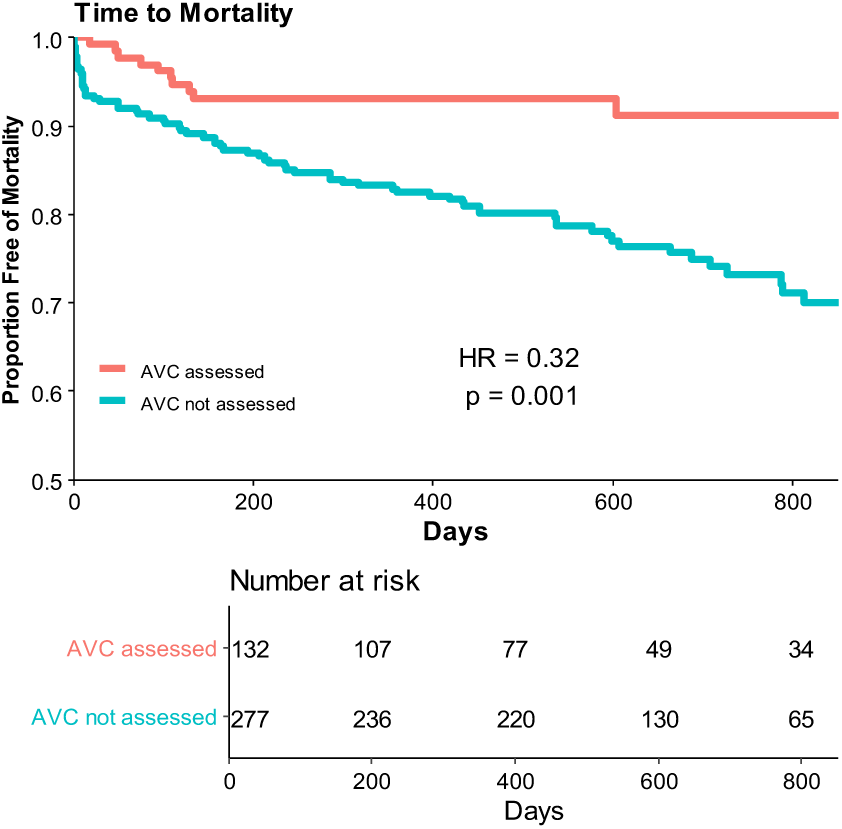
Kaplan-Meier Analysis of All-Cause Mortality by Aortic Valve Calcium (AVC) Assessment Status Kaplan-Meier curves showing time to all-cause mortality in patients with moderate aortic stenosis who underwent AVC scoring (red line) versus those who did not (blue line). Patients who received AVC assessment had significantly lower all-cause mortality compared to those who did not (hazard ratio [HR] = 0.32; p = 0.001). The number at risk at each time point is shown below the plot.

In both the CT cohort and the no-CT cohort, most of the patients who underwent AVR reached a diagnosis of severe AS on follow up imaging prior to the valve intervention (62.8% and 70.3%, respectively). There were 7 patients who underwent surgical AVR with either concomitant CABG or additional valve surgery, or ascending aortic repair in the CT cohort, whereas there were 6 patients in the no-CT cohort who underwent surgical AVR with concomitant CABG or additional valve surgery. These patients were diagnosed as having MAS prior to the surgical procedure. There was one patient in the CT-cohort and two patients in the no-CT cohort who underwent intraoperative TEE with the plan to proceed with TAVR if severe AS were diagnosed. The diagnosis of MAS was revised to severe AS after a comprehensive review of multimodality imaging data in a Heart Team meeting in three patients in the CT-cohort and two patients in the no-CT cohort. The diagnosis of MAS was revised to LFLG-AS after a comprehensive review of multimodality imaging data in a Heart Team meeting in four patients in the CT-cohort and one patient in the no-CT cohort.

All-cause mortality was significantly reduced (HR 0.32, CI 0.16-0.61, p=0.001) in the CT cohort, compared to the non-CT cohort (Figure 3). Median time to all-cause mortality was not statistically significantly different between the two cohorts (101 days vs. 167 days, p=0.418). A significantly higher number of patients died in the no-CT cohort compared with the CT cohort (24.9% vs. 7.6%, p<0.001). There were 22 deaths due to cardiovascular disease (CVD) causes in the no-CT cohort compared to 5 in the CT cohort (7.9% vs. 3.8%, p=0.171). Additionally, there was a significantly higher number of non-CVD related deaths in the no-CT cohort with 47 patients, compared to 5 patients in the CT cohort (17.0%. vs. 3.8%, p<0.001).

### 3.3. Secondary outcomes

Follow-up imaging procedures included repeat TTE, left heart catheterization with dobutamine stress challenge, transesophageal echocardiography (TEE), and low dose dobutamine stress echocardiography. Repeat TTE was the most common imaging modality for follow up imaging, though significantly more common in the no-CT cohort compared to the CT cohort (59.2% vs. 45.5%, p=0.012). In contrast, TEE was performed more often in the CT cohort (28.8%) than in the no-CT cohort (10.1%, p<0.001). Use of dobutamine-based stress testing was infrequent overall, with low absolute numbers in both cohorts (Table 2).

**Table 2.**
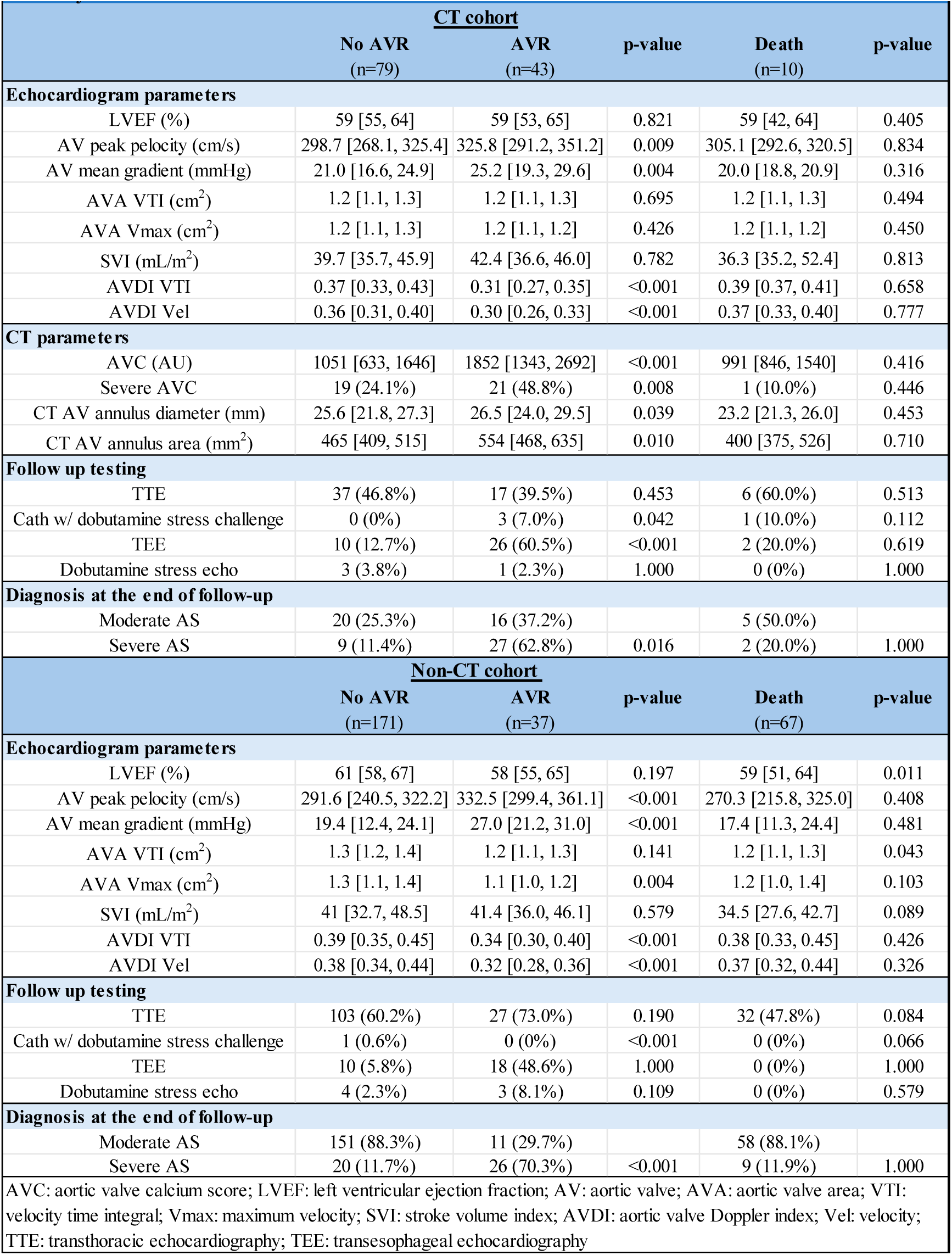
Subgroup Analysis of Baseline Echo Findings, Follow-Up Testing, and Final Diagnoses by AVR and Mortality in the CT and no-CT cohorts.

| **Table 2. Subgroup Analysis of Baseline Echo Findings, Follow-Up Testing, and Final Diagnoses by AVR and Mortality** |  |  |  |  |  |
| --- | --- | --- | --- | --- | --- |
| **CT cohort** |  |  |  |  |  |
|  | **No AVR**<br>**(n=79)** | **AVR**<br>**(n=43)** | **p-value** | **Death**<br>**(n=10)** | **p-value** |
| **Echocardiogram parameters** |  |  |  |  |  |
| LVEF (%) | 59 [55, 64] | 59 [53, 65] | 0.821 | 59 [42, 64] | 0.405 |
| AV peak pelocity (cm/s) | 298.7 [268.1, 325.4] | 325.8 [291.2, 351.2] | 0.009 | 305.1 [292.6, 320.5] | 0.834 |
| AV mean gradient (mmHg) | 21.0 [16.6, 24.9] | 25.2 [19.3, 29.6] | 0.004 | 20.0 [18.8, 20.9] | 0.316 |
| AVA VTI (cm<sup>2</sup>) | 1.2 [1.1, 1.3] | 1.2 [1.1, 1.3] | 0.695 | 1.2 [1.1, 1.3] | 0.494 |
| AVA Vmax (cm<sup>2</sup>) | 1.2 [1.1, 1.3] | 1.2 [1.1, 1.2] | 0.426 | 1.2 [1.1, 1.2] | 0.450 |
| SVI (mL/m<sup>2</sup>) | 39.7 [35.7, 45.9] | 42.4 [36.6, 46.0] | 0.782 | 36.3 [35.2, 52.4] | 0.813 |
| AVDI VTI | 0.37 [0.33, 0.43] | 0.31 [0.27, 0.35] | <0.001 | 0.39 [0.37, 0.41] | 0.658 |
| AVDI Vel | 0.36 [0.31, 0.40] | 0.30 [0.26, 0.33] | <0.001 | 0.37 [0.33, 0.40] | 0.777 |
| **CT parameters** |  |  |  |  |  |
| AVC (AU) | 1051 [633, 1646] | 1852 [1343, 2692] | <0.001 | 991 [846, 1540] | 0.416 |
| Severe AVC | 19 (24.1%) | 21 (48.8%) | 0.008 | 1 (10.0%) | 0.446 |
| CT AV annulus diameter (mm) | 25.6 [21.8, 27.3] | 26.5 [24.0, 29.5] | 0.039 | 23.2 [21.3, 26.0] | 0.453 |
| CT AV annulus area (mm<sup>2</sup>) | 465 [409, 515] | 554 [468, 635] | 0.010 | 400 [375, 526] | 0.710 |
| **Follow up testing** |  |  |  |  |  |
| TTE | 37 (46.8%) | 17 (39.5%) | 0.453 | 6 (60.0%) | 0.513 |
| Cath w/ dobutamine stress challenge | 0 (0%) | 3 (7.0%) | 0.042 | 1 (10.0%) | 0.112 |
| TEE | 10 (12.7%) | 26 (60.5%) | <0.001 | 2 (20.0%) | 0.619 |
| Dobutamine stress echo | 3 (3.8%) | 1 (2.3%) | 1.000 | 0 (0%) | 1.000 |
| **Diagnosis at the end of follow-up** |  |  |  |  |  |
| Moderate AS | 20 (25.3%) | 16 (37.2%) |  | 5 (50.0%) |  |
| Severe AS | 9 (11.4%) | 27 (62.8%) | 0.016 | 2 (20.0%) | 1.000 |
| **Non-CT cohort** |  |  |  |  |  |
|  | **No AVR**<br>**(n=171)** | **AVR**<br>**(n=37)** | **p-value** | **Death**<br>**(n=67)** | **p-value** |
| **Echocardiogram parameters** |  |  |  |  |  |
| LVEF (%) | 61 [58, 67] | 58 [55, 65] | 0.197 | 59 [51, 64] | 0.011 |
| AV peak pelocity (cm/s) | 291.6 [240.5, 322.2] | 332.5 [299.4, 361.1] | <0.001 | 270.3 [215.8, 325.0] | 0.408 |
| AV mean gradient (mmHg) | 19.4 [12.4, 24.1] | 27.0 [21.2, 31.0] | <0.001 | 17.4 [11.3, 24.4] | 0.481 |
| AVA VTI (cm<sup>2</sup>) | 1.3 [1.2, 1.4] | 1.2 [1.1, 1.3] | 0.141 | 1.2 [1.1, 1.3] | 0.043 |
| AVA Vmax (cm<sup>2</sup>) | 1.3 [1.1, 1.4] | 1.1 [1.0, 1.2] | 0.004 | 1.2 [1.0, 1.4] | 0.103 |
| SVI (mL/m<sup>2</sup>) | 41 [32.7, 48.5] | 41.4 [36.0, 46.1] | 0.579 | 34.5 [27.6, 42.7] | 0.089 |
| AVDI VTI | 0.39 [0.35, 0.45] | 0.34 [0.30, 0.40] | <0.001 | 0.38 [0.33, 0.45] | 0.426 |
| AVDI Vel | 0.38 [0.34, 0.44] | 0.32 [0.28, 0.36] | <0.001 | 0.37 [0.32, 0.44] | 0.326 |
| **Follow up testing** |  |  |  |  |  |
| TTE | 103 (60.2%) | 27 (73.0%) | 0.190 | 32 (47.8%) | 0.084 |
| Cath w/ dobutamine stress challenge | 1 (0.6%) | 0 (0%) | <0.001 | 0 (0%) | 0.066 |
| TEE | 10 (5.8%) | 18 (48.6%) | 1.000 | 0 (0%) | 1.000 |
| Dobutamine stress echo | 4 (2.3%) | 3 (8.1%) | 0.109 | 0 (0%) | 0.579 |
| **Diagnosis at the end of follow-up** |  |  |  |  |  |
| Moderate AS | 151 (88.3%) | 11 (29.7%) |  | 58 (88.1%) |  |
| Severe AS | 20 (11.7%) | 26 (70.3%) | <0.001 | 9 (11.9%) | 1.000 |
| AVC: aortic valve calcium score; LVEF: left ventricular ejection fraction; AV: aortic valve; AVA: aortic valve area; VTI: velocity time integral; Vmax: maximum velocity; SVI: stroke volume index; AVDI: aortic valve Doppler index; Vel: velocity; TTE: transthoracic echocardiography; TEE: transesophageal echocardiography |  |  |  |  |  |

In the full CT cohort (n=132), 38 patients (28.8%) received a final diagnosis of severe AS, while in the no-CT cohort (n=277), 55 patients (19.9%) were diagnosed with severe AS during follow-up (p=0.037). Furthermore, the median time to diagnosis of severe AS was significantly shorter in the CT cohort compared to the no-CT cohort (93 days [IQR, 33.89-193.9] vs. 310 days [IQR, 179-472], p<0.001). Median time to AVR was significantly reduced for the CT cohort compared to the no-CT cohort (97 days vs. 394 days, p <0.001) (table 3).

**Table 3.** Median time to valve intervention, mortality and diagnosis of severe aortic stenosis.

| **Table 3. Time to AVR, Mortality, Severe AS Diagnosis, and Follow-Up by AVC Assessment Status** |  |  |  |  |  |
| --- | --- | --- | --- | --- | --- |
|  | **CT cohort** |  | **Non-CT cohort** |  | **p-value** |
|  | **n** | **Days, median [IQR]** | **n** | **Days, median [IQR]** |  |
| Time to AVR | 43 | 97 [63, 165] | 39 | 394 [267-626] | <0.001 |
| Time to mortality | 10 | 101 [55,124] | 69 | 167 [14-435] | 0.418 |
| Time to severe AS diagnosis | 38 | 93 [34,194] | 55 | 310 [179-472] | 0.001 |
| Follow up time | 132 | 492 [256, 822] | 277 | 582 [443-788] | 0.104 |

## 4. Discussion

In this retrospective, single-center cohort study of patients with MAS, we evaluated the clinical utility of integrating computed tomography derived AVC scoring into routine diagnostic workflows. Our primary findings indicate that patients who underwent AVC scoring were diagnosed with severe AS more frequently and underwent aortic valve replacement (AVR) significantly earlier than patients who did not undergo AVC evaluation. Furthermore, all-cause mortality was significantly lower in the CT cohort despite comparable baseline characteristics, underscoring the potential of AVC scoring to refine risk stratification and improve outcomes in this intermediate-risk population.

This study supports the growing recognition that MAS is not a benign condition. Mounting evidence suggests that patients with MAS may experience progressive symptoms, adverse remodeling, and even death in the absence of timely intervention. ^[10]^ However, the diagnosis of MAS remains challenging, particularly when traditional echocardiographic parameters are discordant. ^[11]^ Factors such as suboptimal image quality, inter-operator variability in left ventricular outflow tract (LVOT) measurement, and flow-dependent gradients contribute to diagnostic ambiguity. ^[11]^ AVC scoring, by providing a reproducible and flow-independent quantitative assessment of calcific burden, offers a solution to these challenges and enhances diagnostic clarity. ^[11]^

Our findings are consistent with prior studies showing that AVC burden correlates strongly with AS severity and prognosis. ^[12,13]^ Clavel et al. established that AVC burden strongly correlates with AS severity and can help differentiate between concordant and discordant grading scenarios. ^[13]^ Prior large-scale studies have demonstrated that severe AVC is independently associated with excess mortality, and that AVR improves survival in these patients. ^[12]^ In our study, patients in the CT cohort not only had a higher likelihood of receiving a definitive diagnosis of severe AS but also underwent AVR more promptly (median time to AVR 97 days vs. 394 days). Importantly, the rate of severe AVC burden in the CT group was 33.6%, highlighting that a substantial proportion of patients classified as MAS by echocardiography alone may, in fact, harbor advanced disease.

The clinical implications of these findings are significant. Patients with MAS and elevated AVC may represent a subgroup at heightened risk for adverse events, meriting closer surveillance and potentially earlier intervention. By facilitating the reclassification of disease severity and expediting referral to structural heart teams, AVC scoring may shorten diagnostic delays and improve survival. Our data suggest that mortality was significantly reduced in the CT cohort (HR 0.32, 95% CI 0.16–0.61, p=0.001), and this benefit persisted despite similar observation periods and comparable comorbidity burdens across groups.

The mechanism underlying this mortality reduction is likely multifactorial. First, AVC scoring may improve diagnostic certainty, prompting more timely and targeted intervention. Second, the CT cohort underwent a higher rate of follow-up imaging (including TEE and repeat TTE) and heart team discussions, suggesting a more proactive diagnostic and management approach.

Third, integration of AVC into care pathways may have enhanced shared decision-making between clinicians and patients, increasing the likelihood of appropriate referral for AVR in patients with ambiguous or borderline findings on echocardiography. Finally, there was an excess of non-cardiovascular mortality in the no-CT cohort. It is possible that these patients may be sicker from non-cardiovascular comorbidities.

Aortic stenosis is a complex condition characterized by variable hemodynamic profiles. Accurate classification of severity requires integration of multiple parameters, including aortic valve area (AVA), mean gradient, and flow indices. However, discordant grading can arise from several factors, such as measurement variability or coexisting conditions like hypertension and reduced arterial compliance. ^[13,14]^ Errors in echocardiographic assessment—such as inaccurate left ventricular outflow tract (LVOT) measurements or suboptimal Doppler alignment—can also contribute to diagnostic uncertainty. In these settings, AVC scoring provides a reproducible and objective measure of disease burden and may serve as a useful adjunct in refining the diagnosis. Several additional biomarkers and imaging parameters have been proposed for risk stratification in MAS, including global longitudinal strain, NT-proBNP, high-sensitivity troponin I, and lipoprotein(a). ^[3–5, 15–17]^ However, AVC scoring remains uniquely valuable for its ability to directly quantify valve calcification and aid in distinguishing truly severe disease in cases where TTE findings are ambiguous.

Despite these insights, the optimal timing for intervention in patients with MAS remains unclear. To date, no randomized controlled trials have demonstrated whether earlier valve replacement based on AVC scoring improves clinical outcomes in this population. Ongoing trials such as PROGRESS (NCT04889872), TAVR UNLOAD (NCT02661451), and Evolut EXPAND TAVR II (NCT05149755) are expected to provide more definitive evidence regarding the benefits of early intervention in patients with MAS and elevated risk profiles.

This study has several limitations that should be considered. First, the decision to pursue AVC scoring was made at the discretion of the treating physician, introducing potential referral and selection bias. The worse AV hemodynamics noted in the CT-cohort may be one of the factors that favored AVC assessment for these patients. This variability may reflect differences in provider specialty, experience, or clinical judgment. Although baseline characteristics were largely comparable between the CT and no-CT cohorts, the non-randomized, observational design limits the ability to draw causal inferences.

Second, the analysis was limited by the use of dichotomous variables for AVC scoring, based on previously established sex-specific thresholds for severe calcification. This approach restricted a more granular analysis of the relationship between the degree of calcification and clinical outcomes. Continuous assessment of AVC burden may provide additional insights into risk stratification and outcome prediction, however, is not currently utilized clinically.

Third, the retrospective design inherently limits control over confounding variables. Additionally, the timing and modality of follow-up imaging and intervention were not standardized, which may have influenced clinical decision-making and outcome ascertainment. Due to these limitations, it is not possible to establish a direct cause-effect relationship between AVC scoring and clinical outcomes based on this study alone. However, the findings support the hypothesis that AVC scoring may play a valuable role in guiding earlier intervention, and they provide a foundation for future prospective studies to explore this relationship more definitively.

Our findings support the use of AVC scoring to enhance diagnostic clarity and inform clinical decision-making in patients with MAS. Future prospective studies are needed to determine whether high AVC burden alone—irrespective of standard echocardiographic findings—should prompt closer surveillance or consideration of early AVR in patients with concordant MAS. Identifying the subset of patients with MAS who are most likely to benefit from earlier intervention remains a critical research priority. In particular, studies should explore whether AVC scoring can serve as a standalone marker to guide timing of AVR and define optimal AVC cut-off values for clinical decision-making. Additionally, integration of AVC with other risk markers—such as biomarkers, functional assessment, and myocardial imaging—may further refine individualized care strategies in moderate AS. Randomized controlled trials are essential to determine whether earlier AVR in patients with high AVC burden translates into improved survival, symptom burden, and quality of life.

## 5. Conclusion

AVC scoring is a valuable tool for risk stratification and for identifying high-risk patients with MAS who may benefit from earlier valve intervention. Our study supports the use of CT-derived AVC as a prognostic imaging marker that can be effectively integrated into routine clinical practice as a complementary modality alongside echocardiography. Incorporating CT-AVC into the diagnostic and surveillance strategy for patients with MAS may enhance risk assessment, inform management decisions, and facilitate timely referral for valve replacement in appropriate candidates.

## Data Availability

Data will be made available upon request

## Acknowledgements

None

## Tables and Figure Legends

**Central Illustration:**
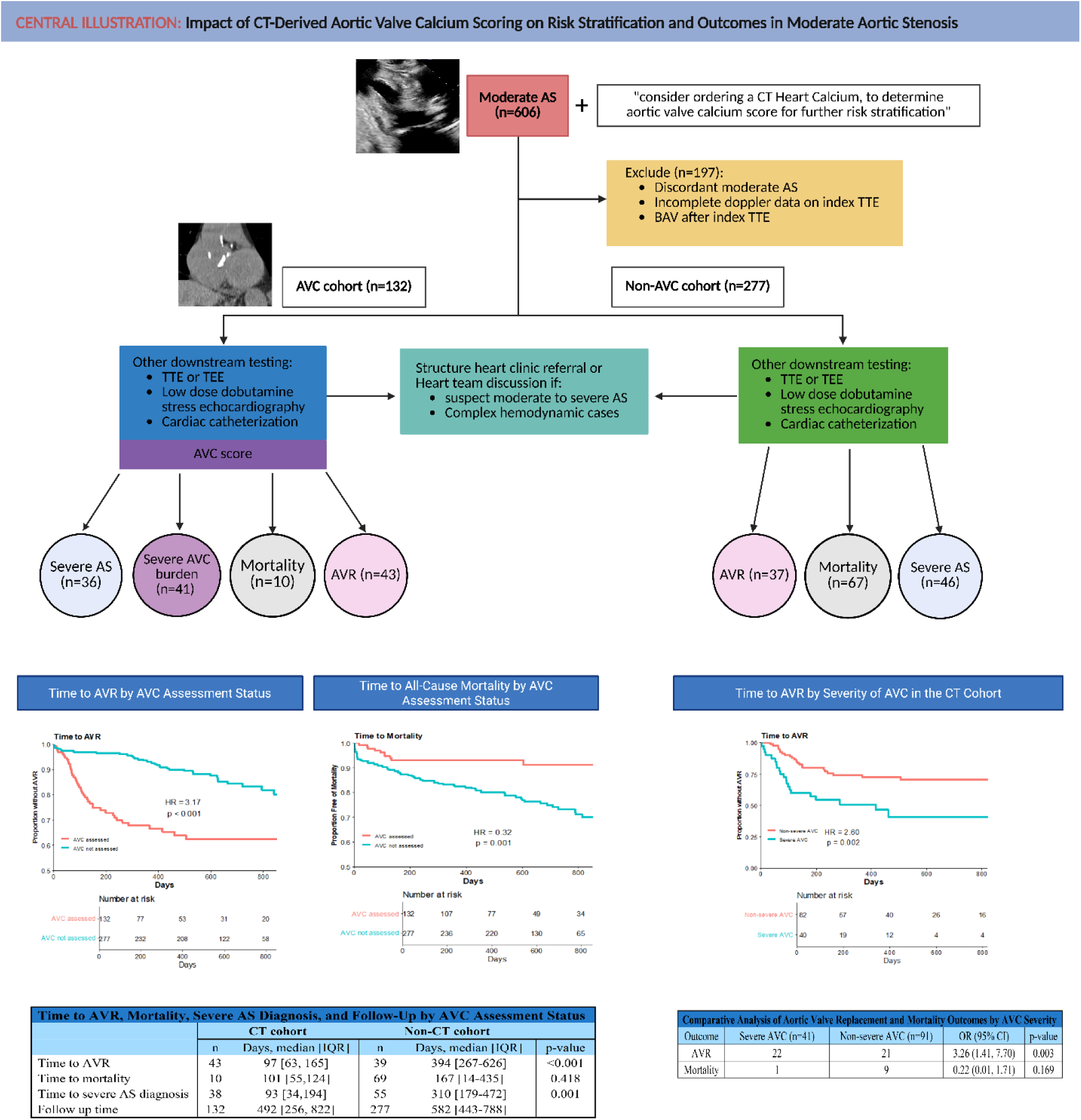
Figure shows prevalence of severe aortic stenosis, median times for diagnosis of severe aortic stenosis, all-cause mortality and aortic valve intervention in the non-CT vs. CT-AVC cohorts. Kaplan Meier curve results for valve intervention for CT and no-CT cohorts and patients with non-severe vs. severe aortic valve calcification in the CT cohort.

